# Antimicrobial Resistance Gap Analysis for Uropathogens in Nigeria: Pathogen Profiles, Resistance Burden, and Translational Microbiological Action Plans

**DOI:** 10.1101/2025.07.27.25332264

**Authors:** Nooruldeen Saad, Jafar Eyad, Amjad B.A Banibella, Ramadhani Chambuso

## Abstract

**Background:** We investigated laboratory culture results for uropathogens with existing antimicrobial resistance (AMR) patterns to leverage novel microbiological gaps in Nigeria.

**Methods:** In a retrospective study, we analysed a total of 84,548 valid culture results from 26,630 patients across 25 public laboratories participated in the national AMR report. The WHO priority bacteria uropathogens with their AMR patterns for the key urinary tract infections (UTIs) antibiotic classes were assessed. A p-value of **<0.05** was considered statistically significant.

**Results:** Urine contributed 6426 samples collected for culture. A total of 16 uropathogens were identified, *Escherichia coli* was the most frequently isolated organism each year. Among Gram-negatives, *Klebsiella species* and *Escherichia coli* showed alarming resistance to cephalosporins (80% each). *Staphylococcus aureus* demonstrated the highest resistance burden, with methicillin and folate pathway inhibitor resistance reaching 89% and 82%, respectively. In the gap analysis, cephalosporin susceptibility testing remained incomplete and in the mapping of six uropathogens and their recommended stewardship responses, *E.coli* was recommended to alert the monitoring of tetracycline and folate inhibitors.

**Conclusion:** High AMR across key drug classes for uropathogens like *Klebsiella, Escherichia* and *Staphylococcus species* suggests a narrowing window for effective treatment of UTIs in Nigeria.

**Graphical Abstract:** 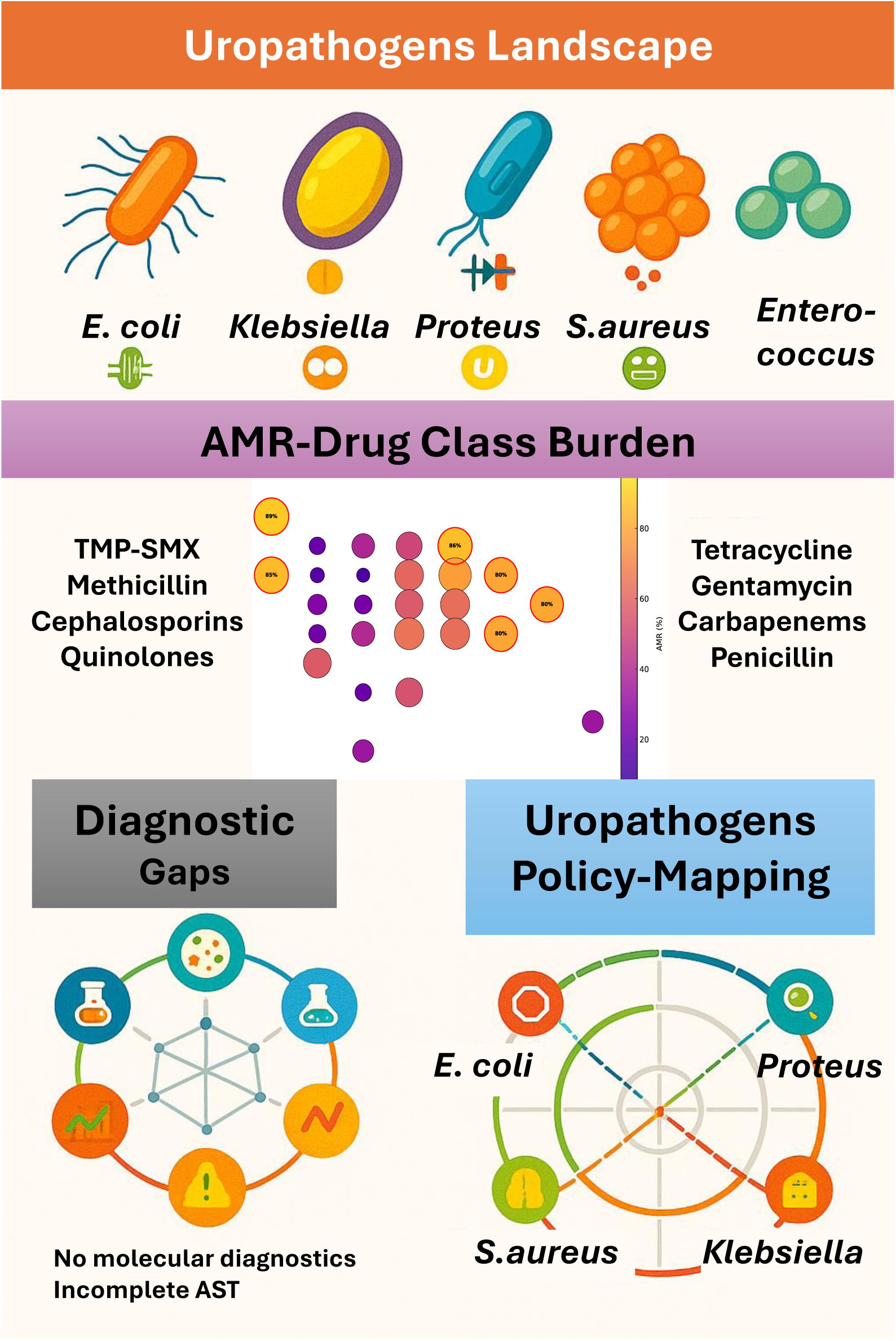

## Introduction

Urinary tract infections (UTIs) caused by uropathogens are among the most common bacterial infections globally, with over 400 million cases annually and significant recurrence rates.[1] In West Africa, the burden of UTIs is underestimated due to fragmented diagnostics and underreporting that UTIs may account for over 30% of hospital acquired infections.[2] In Nigeria, the most populated country in Africa, the local surveillance reports estimate tens of thousands of UTIs each year. UTIs contribute to antimicrobial overuse, hospital readmissions and increased burden of antimicrobial resistance (AMR). AMR has driven repeated empirical treatment failures, especially where diagnostics are limited and treatment faces huge drug resistance.[3] Previous studies have explored pathogen profiles and AMR trends, however a few have integrated uropathogens, drug classes AMR mapping with WHO priority pathogens for UTIs and the microbiological gaps. Most importantly, there are few national-level microbiology-driven gap analysis studies linking AMR data to actionable treatment policy recommendations in Nigeria. [3, 4]

AMR in uropathogens is fundamentally a microbiological phenomenon driven by selective pressure, genetic exchange, diagnostic limitations and therapeutic failure. WHO priority bacteria pathogens for UTIs such as *Escherichia coli*, *Klebsiella pneumoniae*, *Pseudomonas aeruginosa* and *Enterococcus faecium* are frequently the causative agents of UTIs, especially in healthcare-associated infections.[5] These organisms exhibit complex resistance mechanisms including extended-spectrum β-lactamase (ESBL) production, carbapenemase activity, and biofilm formation. For example *E. coli*, the most common uropathogen produces fimbriae with which it sticks to the wall of the urinary bladder to produce toxins that destroy bladder cells.[6, 7] Also, *Klebsiella species* have a capsular protective coating layer that shields itself from the host immune system and enables the pathogen to be antibiotic-resistant, especially with beta-lactams.[8, 9] The increasing resistance of uropathogens to first-line antimicrobial treatment compromises UTIs therapy and lead to prolonged infection control challenges. This promotes the spread of resistant strains and complicates the management of UTIs which is now a global health problem.[10–12]

We hypothesize that specific uropathogens, particularly the WHO priority Gram-negatives have high AMR thresholds across the key UTIs antibiotic classes, yet this remains unlinked to treatment guidelines and health policy action plans for UTIs in Nigeria.[13, 14] This gap between laboratory resistance data and empirical treatment practices reflects a critical microbiological disconnect that undermines the effectiveness of AMR stewardship in lower middle-income countries.

In this gap analysis study, we analyzed existing national AMR report with laboratory culture results for uropathogens and the existing AMR patterns to leverage novel microbiological insights that can guide national-level evidence-based interventions for UTIs management in Nigeria.

## Methodology

### Study design

A retrospective study of the national AMR surveillance report from Nigeria for the years 2016 to 2018, similar to a published study. [15] This published report was generated by the Mapping Antimicrobial Resistance and Antimicrobial Use Partnership (MAAP) consortium. It involved 25 sentinel laboratories across Nigeria with bacteriology testing capacity. This study adheres to the STROBE (Strengthening the Reporting of Observational Studies in Epidemiology) guidelines.[16]

### Data sources and surveillance framework

The original data were collected by Nigeria health authorities in collaboration with MAAP partners.[17] Surveillance activities were carried out across a national network of medical laboratories selected for their bacteriology testing capacity. A total of 25 laboratories contributed antimicrobial susceptibility testing (AST) data using WHONET software, a standardized microbiology data management tool.[18] Data collection involved trained field teams retrieving laboratory records from both paper-based and digital systems. Where feasible, these records were linked with hospital databases to include patient demographics and clinical metadata (e.g., age, sex, specimen type). The surveillance focused on human health and isolates were obtained from urine and other clinical specimens. The uropathogens reported from culture results include:

a. Positive cultures with no specie identification

(i) Klebsiella species
(ii) Proteus species
(iii) Pseudomonas species
(iv) Enterococcus species
(v) Staphylococcus species
b. Positive cultures with specie identification

(i) Klebsiella aerogenes
(ii) *Klebsiella oxytoca*
(iii) *Klebsiella pneumoniae*
(iv) *Escherichia coli*
(v) *Escherichia fergusonii*
(vi) *Proteus hauseri*
(vii) *Proteus mirabilis*
(viii) *Proteus vulgaris*
(ix) *Pseudomonas aeruginosa*
(x) *Pseudomonas fluorescen*
(xi) *Pseudomonas stutzeri*
(xii) *Enterococcus faecalis*
(xiii) *Staphylococcus aureus*

### Inclusion criteria and data Management

Only isolates from the above bacterial groups with valid AST results were included in the analysis. In accordance with WHO Global Antimicrobial Resistance Surveillance System (GLASS) guidelines and Clinical and Laboratory Standards Institute (CLSI) M39-A4 recommendations, only the first isolate per patient per year was retained to avoid duplicate counting.[19, 20] In cases where unique patient identifiers were not available, all isolates were retained and interpreted with appropriate caution. AST interpretations followed CLSI criteria, and where necessary, zone diameters or minimum inhibitory concentrations (MICs) were standardized using WHONET’s interpretive rules to ensure consistency across laboratories.[21, 22]

### Data extraction from the report

In this study, we manually extracted relevant pathogen-specific data from the national report, including:

(i) Total number of participating laboratories
(ii) Collected specimens per year
(iii) Source of collected specimens
(iv) Specimen types
(v) Valid cultures
(vi) Positive and negative cultures
(vii) Species-level breakdown (e.g., *E. coli, K. pneumoniae, P. aeruginosa*, etc.)
(viii) AST results for antibiotics tested
(ix) Patient demographics (age and sex)

These variables were compiled into a structured Microsoft Excel dataset with multiple spreadsheets for different variables (**Supplementary Datasets**) to enable consistent data analysis. Species were identified where possible, while others were recorded generically as *Klebsiella spp., Escherichia spp., Proteus spp., Pseudomonas spp.*, or *Enterococcus spp*., according to how they appeared in the original report. AMR data were available for multiple antibiotics from antibiograms and figures for the analysis of year-wise resistance trends and identification of the most prevalent resistant antibiotic combinations.

### AST and quality control

AST was conducted using both disk diffusion and MIC panels, depending on each laboratory’s capacity. The primary interpretive standard applied was CLSI criteria. Quality control strains were widely used for routine AST quality control. External Quality Assessment (EQA) was implemented across participating laboratories through a structured proficiency testing program coordinated by MAAP and the Fleming Fund Laboratories were encouraged to participate in periodic EQA rounds, and corrective actions were documented for laboratories with substandard performance.[23, 24] However, details on the proportion of laboratories consistently participating in EQA or their inter-laboratory performance were not systematically reported.

### AMR calculation

From the original report, AMR rates were derived from positive cultures with available AST results. AMR rates were calculated as the proportion of non-susceptible isolates (intermediate or resistant) relative to the total number of tested isolates within a single calendar year.

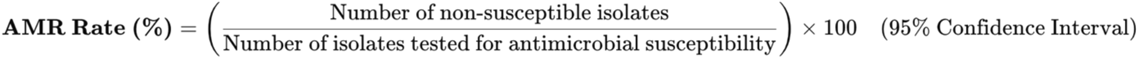

Where:

- Non-susceptible isolates = Resistant + Intermediate
- Tested isolates = All isolates subjected to AST

### Statistical analysis

Statistical analyses and figure visualizations were peformed using R version 4.3.2 and Python version 3.11. Missing data were about <1% and handled via complete-case analysis. We analysed valid culture results using descriptive statistics to compare culture positivity across demographic groups and years. Annual culture positivity rates were calculated with 95% Wald confidence intervals (CI). The heterogeneity across years was evaluated using Chi-square test. For categorical comparisons (e.g., sex, age group, year), the proportions for culture positivity were calculated using Pearson’s Chi-square test. A p-value of **<0.05** was considered statistically significant.[25, 26]

## Results

### Demographics and culture characteristics

Among 84,548 valid cultures analysed, the majority were obtained from female patients (55.6%), who also exhibited a significantly higher culture positivity rate compared to males (35.1% vs 28.3%, respectively (**p<0.001**, **Table 1**). Culture positivity increased steadily with age, reaching 40.0% in those aged >65 years. Although the largest number of cultures originated from adults aged 18–49 years, their positivity rate was moderate (31.8%) compared with older groups. Over the three-year period, culture positivity rose from 28.5% in 2016 to 34.6% in 2018, suggesting either improved diagnostic targeting or a true increase in uropathogens burden.

**Table 1.** Distribution of valid microbiological cultures by gender, age group, and year.

| Variable | Valid cultures, n | Positive cultures, n | Positive cultures with AST, n | Positive cultures (%) | *95% CIs | **p-value |
| --- | --- | --- | --- | --- | --- | --- |
| Gender |  |  |  |  |  |  |
| Male | 37550 | 10618 | 10160 | 28.3 | 27.8 – 28.7 | <0.001 |
| Female | 46997 | 16517 | 13803 | 35.1 | 34.7 – 35.6 |  |
| Age Group |  |  |  |  |  |  |
| < 1 year | 9689 | 2832 | 2711 | 29.2 | 28.3 - 30.1 | <0.001 |
| 1 – 17 (yrs) | 20476 | 4902 | 4526 | 23.9 | 23.4 - 24.5 |  |
| 18 – 49 (yrs) | 29336 | 9323 | 7629 | 31.8 | 31.2 - 32.3 |  |
| 50 – 65 (yrs) | 5177 | 1753 | 1630 | 33.9 | 32.6 - 35.2 |  |
| Above 65 (yrs) | 4743 | 1898 | 1806 | 40.0 | 38.6 - 41.4 |  |
| Unknown age | 15127 | 6427 | 5661 | 42.5 | 41.7 - 43.3 |  |
| Year |  |  |  |  |  |  |
| 2016 | 26896 | 7678 | 6532 | 28.5 | 28.0 - 29.1 | <0.001 |
| 2017 | 32134 | 10640 | 9501 | 33.1 | 32.6 - 33.6 |  |
| 2018 | 25518 | 8817 | 7930 | 34.6 | 34.0 - 35.1 |  |

\*95 % confidence intervals (Wilson method) for culture-positivity % \*\*Chi-Square Test for independence to assess if there was a statistical significant difference in culture positivity across variables

### Diagnostic yield, specimen distribution and temporal trends of uropathogens across surveillance laboratories

Across the 25 sentinel laboratories surveyed, marked heterogeneity was observed in the microbiological diagnostic yield and reporting completeness of cultures (**Fig 1a**). While most laboratories reported a predominance of negative cultures, a concerning proportion of positive cultures lacked antimicrobial susceptibility testing (AST), particularly at Murtala Muhammad, and Minna laboratories. Only a few centres consistently linked culture positivity with AST reporting. Among the top ten leading specimen types, urine was the most frequently processed specimen across all years, peaking in 2018 (**Fig. b)**. A total of 16 uropathogens were identified between 2016 and 2018, ***Escherichia coli*** was the most frequently isolated organism each year (**Fig 1c**). *Klebsiella species* isolates rose from 922 in 2016 to 1245 in 2018. Among species-level identifications, *Klebsiella pneumoniae* peaked in 2017 (851 isolates) before declining in 2018. Other frequently reported pathogens included *Pseudomonas aeruginosa*, *Enterococcus species* and *Proteus mirabilis*, while *Escherichia fergusonii*, *Pseudomonas stutzeri*, and *Proteus hauseri* were rarely detected.

**Fig 1.**
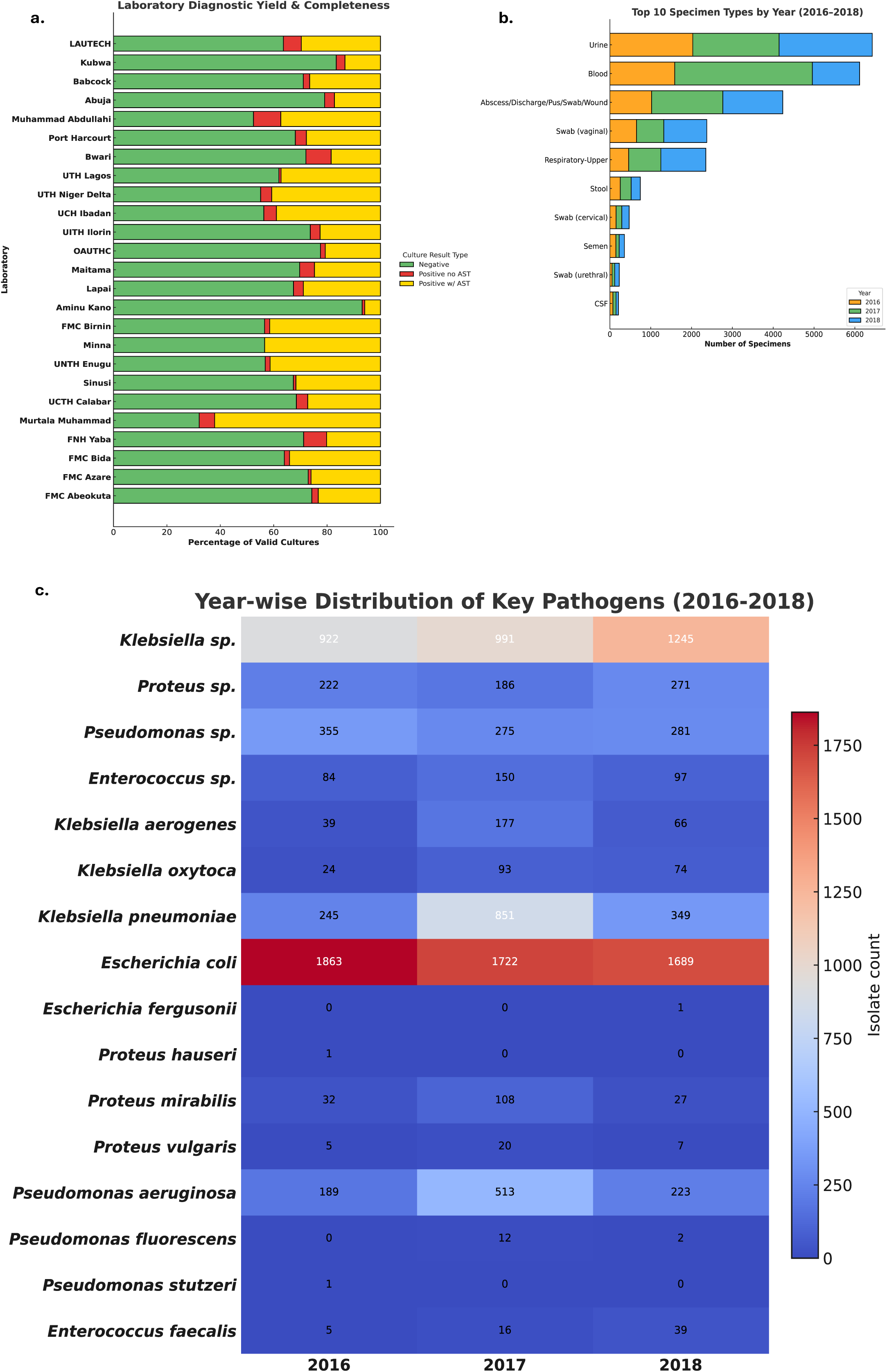
Laboratory diagnostic yield, specimen distribution, and temporal trends. **(a)** Laboratory-level distribution of valid urine cultures categorized by diagnostic yield with inter-laboratory variability, several sites showing high proportions of culture-positive specimens but lacking AST data. **(b)** Bar chart showing the top 10 specimen types processed annually. Urine remained the most common collected specimen type across all years. **(c)** Heatmap of year-wise isolate counts for key uropathogens. *Escherichia coli* consistently ranked as the leading pathogen, with counts declining slightly over the three years.

### Yearly AMR patterns across drug classes and specific uropathogen species

Given the clinical reliance on empiric therapy for uropathogens in low-resource settings, and to understand the AMR patterns across drug classes and bacterial species, we performed a comparative AMR gradient, highlighting the proportion of AMR by pathogen–drug class combinations for year 2016 (**Fig 2**). Among Gram-negatives, *Klebsiella spp*. and *Escherichia coli* showed alarming resistance to cephalosporins (80% each). *Proteus spp.* also exhibited striking resistance to folate pathway inhibitors (91%), a concern for empirical treatment in urinary infections. Interestingly, there was no vancomycin resistance to any pathogens. Notably, *Staphylococcus aureus* and all other *Staphylococcus species* displayed resistance above 80% to methicillin, reinforcing MRSA prevalence.

**Fig 2.**
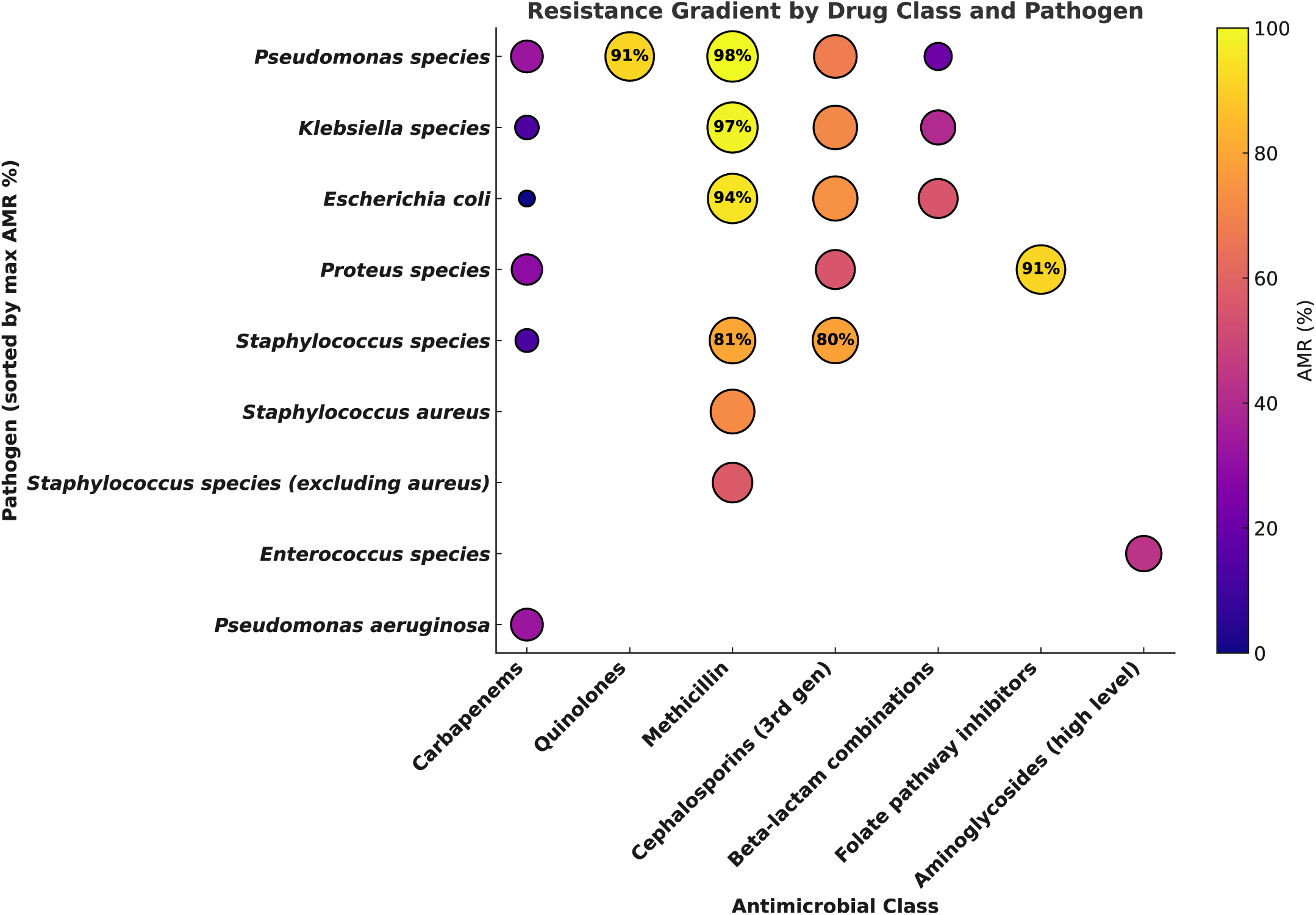
Resistance gradient of priority uropathogens across antimicrobial classes. As expected, extensive resistance to methicillin was observed. *Proteus species* showed 91% resistance to folate pathway inhibitors. Resistance to carbapenems, vancomycin and aminoglycosides remained the lowest across uropathogens. Red-outlined bubbles denote AMR ≥80%, and gradients reflect urgent therapeutic constraints for UTIs. **Note**: Red-bordered bubbles indicate resistance rates ≥80%.

In 2017, *Klebsiella aerogenes* exhibited the highest resistance levels, with 91% to second-generation cephalosporins and 83% to quinolones, reflecting potent β-lactam and fluoroquinolone resistance (**Fig 3**). *Escherichia coli* and *Streptococcus pneumonia* showed elevated resistance to multiple β-lactam classes, including tetracyclines (88% and 81%, respectively), indicating cross-resistance trends. Resistance among *Klebsiella* and *Staphylococcus* species remained moderate but widespread across all tested drug classes, suggesting intrinsic resistance.[27–29]

**Fig 3.**
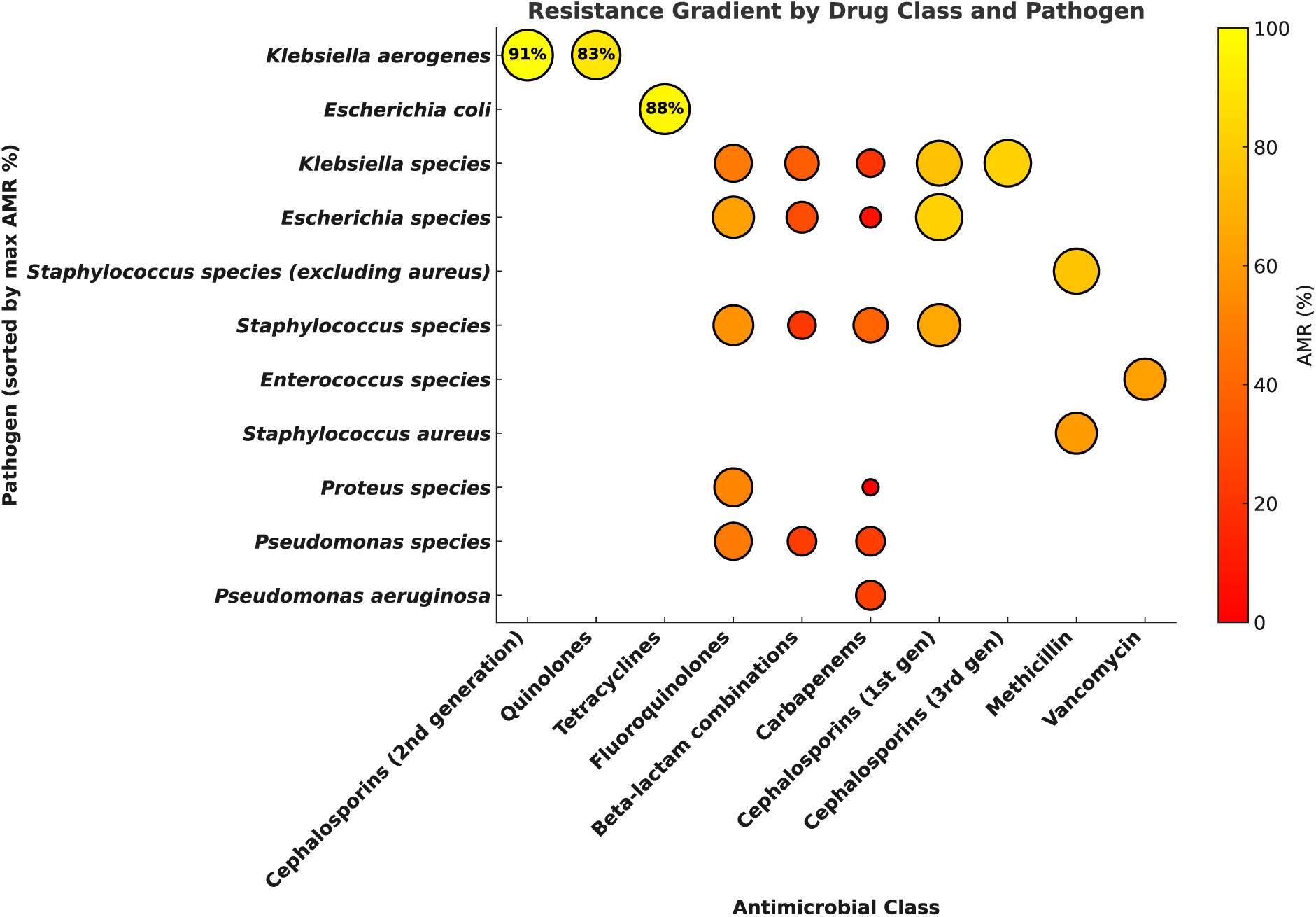
Resistance gradient of priority uropathogens to diverse antimicrobial classes. *Klebsiella aerogenes* showed the most alarming resistance, with 83% and 91% to quinolones and 2nd-generation cephalosporins, respectively. *Escherichia coli* also exhibited high resistance to tetracyclines, reaching 88% AMR in multiple classes. Interestingly, *Pseudomonas aeruginosa* displayed relatively lower AMR rates. **Note**: Black-bordered bubbles indicate resistance rates ≥80%.

In 2018, *Staphylococcus aureus* demonstrated the highest resistance burden, with methicillin and folate pathway inhibitor resistance reaching 89% and 82%, respectively (**Fig 4**). *Pseudomonas species* and *Escherichia coli* also showed striking resistance patterns, particularly to tetracyclines and folate pathway inhibitors (86% and 85%, respectively), highlighting ongoing challenges in treating Gram-negative infections. High resistance to first-generation cephalosporins among *Staphylococcus species* and *Escherichia coli* points to diminishing empirical coverage.

**Fig 4.**
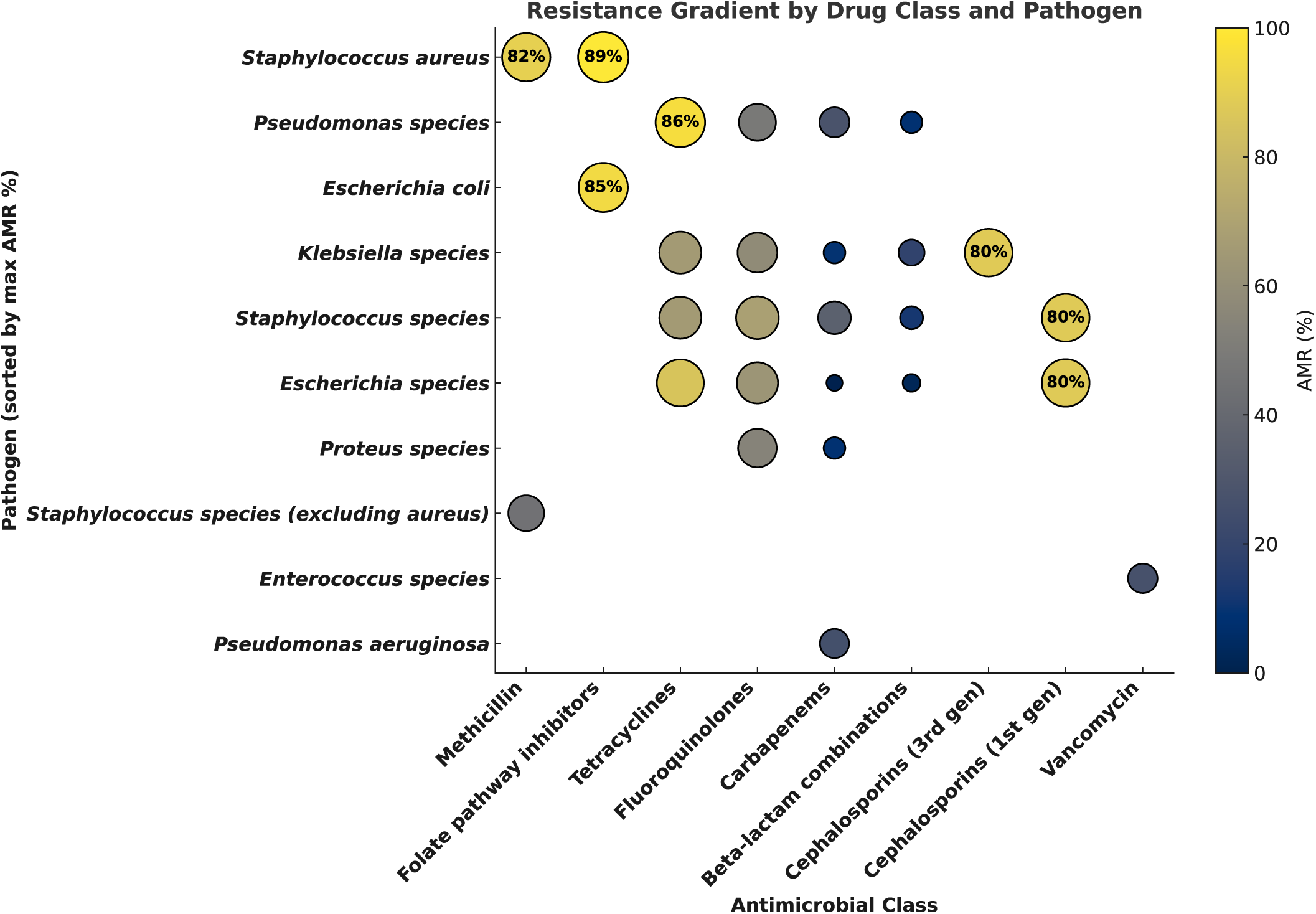
AMR gradients across uropathogens and drug classes. *Staphylococcus aureus* and *Escherichia coli* showed the highest resistance to folate pathway inhibitors (89% and 85%, respectively), to and *Pseudomonas species* demonstrated concerning resistance to tetracycline. Notably, *Klebsiella species* had 80% resistance to third-generation cephalosporins. **Note**: Red-bordered bubbles indicate resistance rates ≥80%.

### AMR analysis for uropathogens to address critical microbiology-related gaps

Understanding AMR gaps in uropathogens is essential for guiding actionable stewardship interventions. We identified six critical microbiology-related gaps that compromise the clinical utility of antibiograms and local resistance mapping (**Fig 5**). Of concern is the widespread absence of molecular AMR gene surveillance and routine ESBL gene panel testing which are both essential for detecting emerging resistance before phenotypic failure. Despite high prevalence, cephalosporin susceptibility testing remains incomplete. Furthermore, the lack of feedback loops to antimicrobial stewardship programme (ASP) teams and the continued reliance on non–data-guided empirical treatment protocols perpetuate inappropriate prescribing.

**Fig 5.**
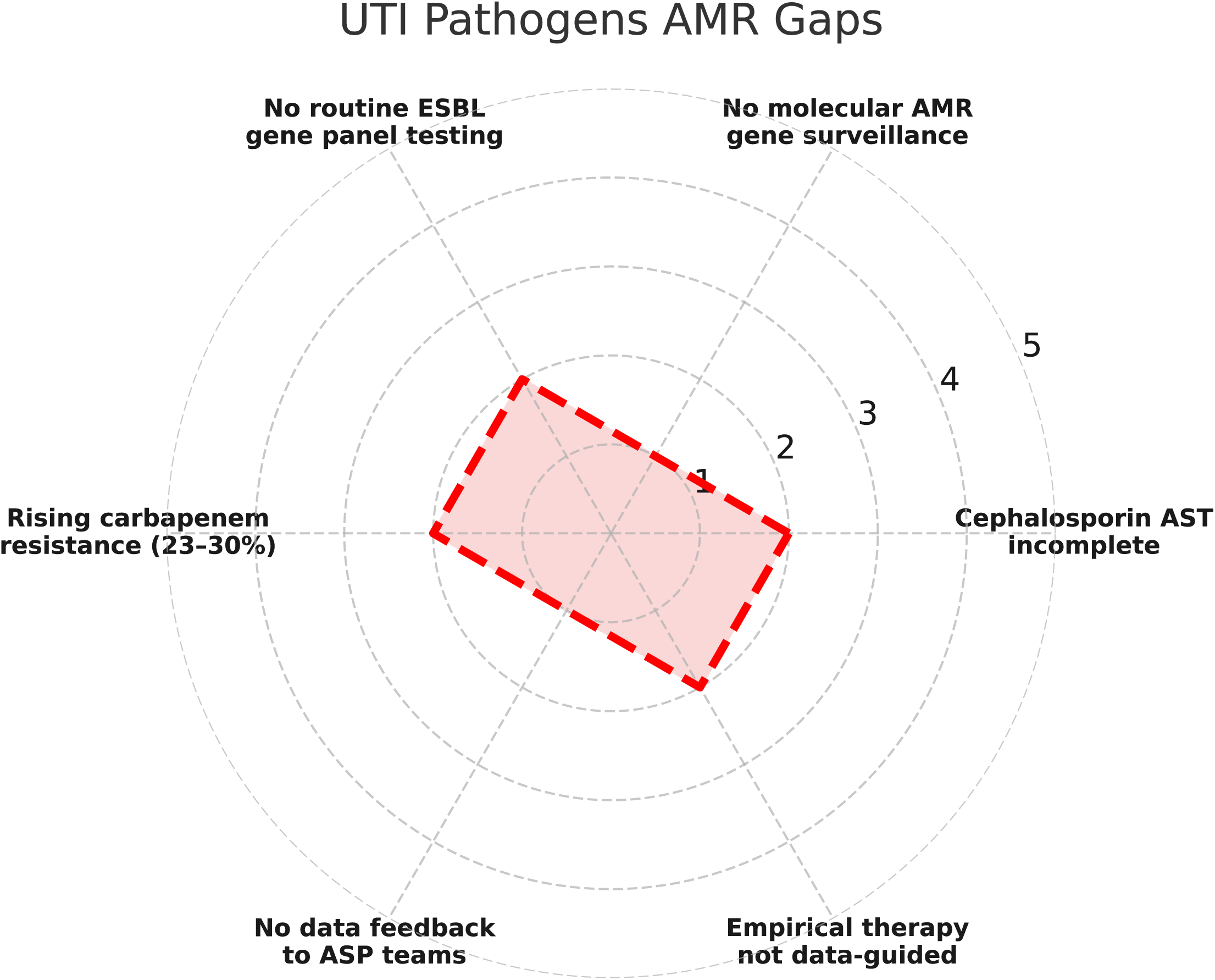
Visualizing AMR gaps in the management of UTIs. Poor diagnostic capacity (e.g., lack of molecular AMR gene surveillance and absence of routine ESBL testing), surveillance deficits (e.g., no feedback loops to antimicrobial stewardship programs), and treatment risks (e.g., rising carbapenem resistance and empiric therapies unaligned with local resistance data) are highlighted. These gaps represent a critical barrier to effective UTIs management, delaying appropriate therapy, increasing resistance selection pressure and undermining infection control efforts.

### Addressing identified pathogen-specific challenges for uropathogens

Understanding how laboratory evidence of AMR translates into actionable clinical policy is a critical gap in surveillance-response frameworks across low-resource settings. Despite increasing availability of pathogen-specific AMR data, translation into stewardship interventions remains fragmented, especially for uropathogens with multidrug resistance profiles. To address that, we performed a structured pathogen–recommendation linkage analysis, visually synthesizing resistance challenges for uropathogens and the targeted recommended policy responses to support more actionable UTIs guidelines (**Fig 6**). This approach offers a novel visual policy–microbiology interface that highlights missed opportunities, redundancy, and priority action points within national AMR plans. A map linking six uropathogens and their recommended stewardship responses show that pathogens with the highest resistance burden such as *Staphylococcus species* (Folate pathway inhibitors 89%, **Fig 4**) were recommended to update the treatment guidelines and perform regional antibiogram alerts and *E.coli* was recommended to alert the monitoring of tetracycline and folate inhibitors. Recurrent linkages, such as multiplex PCR and species-level identification for *Streptococcus* and restrict for cephalosporins, fluoroquinolones and third-generation cephalosporins empirically for *Klebsiella species* reveal diagnostic infrastructure and AMR stewardship gaps.

**Fig 6.**
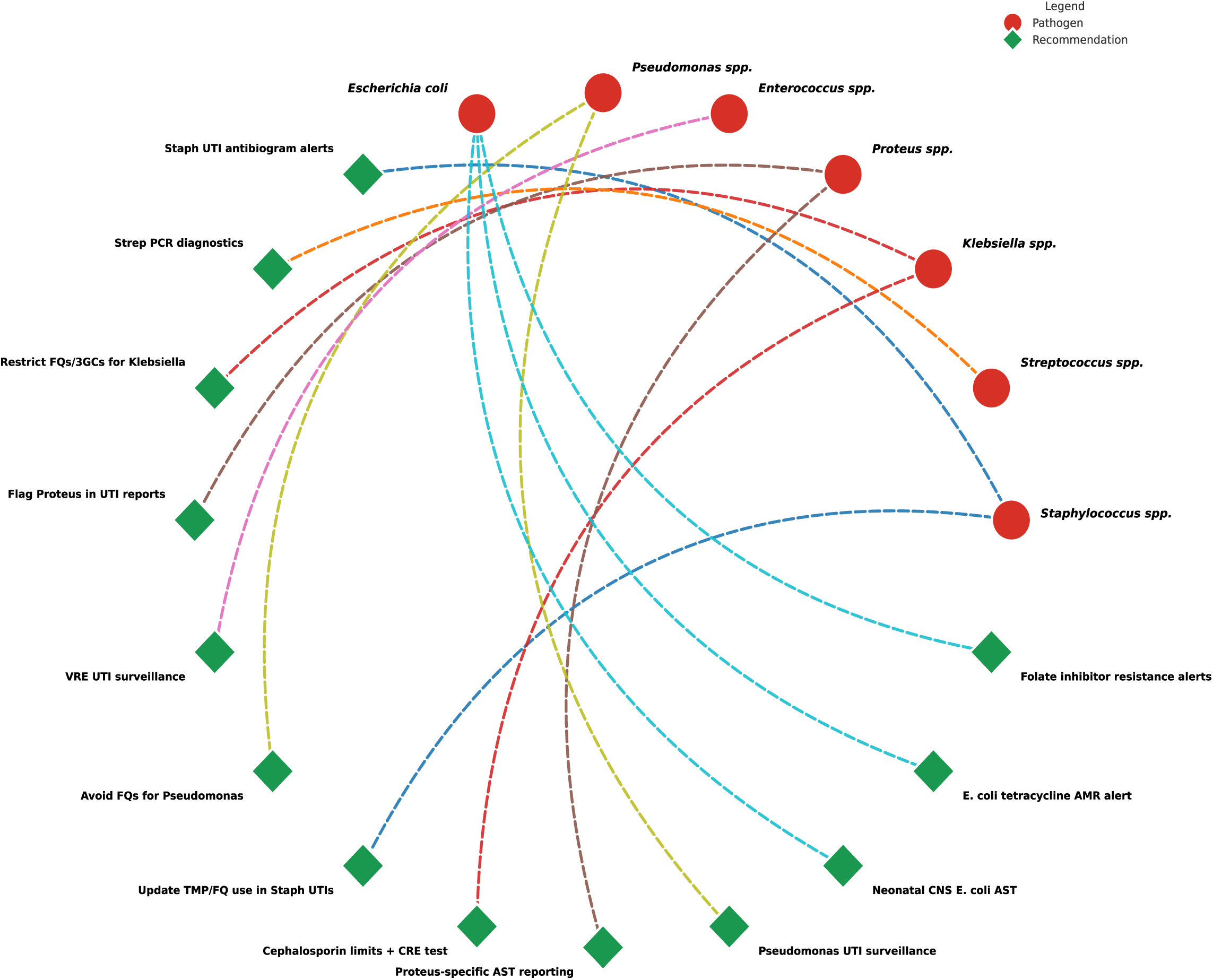
Pathogen-specific AMR recommendations for each uropathogen. Arcs dash lines connect each pathogen to its associated recommendation, with arc colour uniquely mapped to specific pathogen. *E.coli* was recommended to alert the monitoring of tetracycline and folate inhibitors while *Pseudomonas species* recommended to include guidance on excluding fluoroquinolones from empiric protocols and enhancing national surveillance, while *Streptococcus species* were recommended to enhance species identification and multiplex PCR. This mapping supports targeted AMR mitigation and informs evidence-based updates to UTIs treatment policies.

## Discussion

To our knowledge, this is the largest multicentre AMR surveillance study analyzed over 84,000 valid clinical cultures collected from 2016 to 2018 across 25 laboratories in Nigeria and providing a comprehensive gap analysis assessment of uropathogen resistance patterns and microbiology-related gaps. Focusing on the cultures of uropathogens, we systematically mapped annual AMR trends, pathogen-specific burdens and diagnostic stewardship deficits. Also, we provided novel insights include the integration of resistance gradient visualizations, pathogen-to-policy linkage maps and critical microbiological gap analyses tool rarely used in side by side in tandem microbiology studies. Our findings are significant for global health microbiology as they expose the urgent need to align local diagnostic practices and AMR data to reduce inappropriate prescribing and improve UTIs outcomes.

This study reveals critical diagnostic and therapeutic vulnerabilities in uropathogens management in Nigeria. High resistance rates to key UTIs antibiotics, particularly folate pathway inhibitors and cephalosporins, reflect both selective pressure from empirical prescribing and limited access to updated diagnostics.[30, 31] The underutilization of AST and molecular tools compromises the interpretive power of positive cultures led to ineffective treatment decisions and delayed containment of resistant strains. In addition, the disconnect between laboratory performance, AMR burden and clinical policy as evidenced by the absence of actionable stewardship linkages in the gap analysis, highlights a systemic failure in translating laboratory evidence into UTIs treatment guideline updates, reinforcing the importance of integrated AMR surveillance to laboratory policy frameworks.[32, 33]

Findings in this study offer immediate translational potential for strengthening Nigeria’s microbiology-informed health systems response to AMR in uropathogens. By mapping specific pathogen–antibiotic resistance profiles to existing actionable stewardship and recommendations, our study lays the groundwork for evidence-based intervention to UTIs treatment guidelines and antibiogram development in LMICs. The identification of diagnostic gaps, such as incomplete AST and lack of molecular resistance surveillance, emphasizes the urgent priorities for capacity-building in clinical microbiology laboratories.[34, 35] Embedding these insights into the national AMR action plans specifically for UTIs can enhance therapeutic accuracy, support rational antibiotic use, and ultimately reduce resistance selection pressure, aligning Nigeria’s response with global targets for microbiology-driven AMR containment.

The high methicillin resistance among *Staphylococcus species* (>80%) observed in this study was higher than reported in previous UTIs-AMR reports from Nigeria.[36, 37] However, these two studies lacked gap analysis perspective compared to our study which provides a comprehensive 3-year, multi-centre gap analysis perspective with pathogen-level AMR stewardship mapping. Also, this discrepancy could be possibly due to methodological differences or sampling biases. Regional variations in resistance rates may also reflect disparities in laboratory diagnostic capacities, antibiotic use policies or infection control implementation across settings.

Focusing on a global health microbiology perspective, our findings significantly contribute to the understanding of AMR in uropathogens within high-burden regions for UTIs like Nigeria.[38] The alarmingly high resistance rates to frontline antibiotics mirrored trends reported in global surveillance networks like GLASS, emphasising the universal threat posed by resistant uropathogens.[39, 40] However, our data uniquely provide granular, pathogen-specific resistance and stewardship gaps in a West African setting, a region often absent in global meta-analyses for uropathogens and AMR. The study’s structured pathogen–policy mapping approach offers a reproducible model for integrating microbiological evidence into AMR containment strategies in Africa in general, aligning with WHO’s One Health approach framework.[41, 42] These findings not only fill a geographic data void but also provide actionable new insights to inform global AMR policy, diagnostics development, and stewardship interventions for UTIs tailored for low-resource settings.[43]

The strengths of this study include:

(i) Large sample size of total cultures analyzed, providing robust statistical power and generalizability for UTIs cultures.
(ii) Gap analysis study to expose the hidden Pathogen–drug class resistance patterns, allowing actionable clinical interpretation.
(iii) Inclusion of data from three consecutive years (2016–2018) enabled to analyse the trends of AMR evolution in uropathogens.
(iv) Focused on WHO priority bacteria uropathogens, enhancing relevance to global microbiology and AMR surveillance goals.
(v) Identification of diagnostic gaps highlighted critical microbiology weaknesses such as lack of ESBL testing and poor species-level identification of uropathogens.
(vi) Visualization of laboratory/AMR policy with microbiological recommendations, introduce a novel visual mapping of uropathogen specific challenges and the suggested actionable solutions.
(vii) The use of public reports to analyse exist and missing gaps to strengthen treatment outcomes in a global health perspective is innovative.

The potential weaknesses of this study may include:

(i) Our study did not correlate the AMR patterns with patient-level clinical outcomes such as treatment failure, complications or mortality.
(ii) Absence of urine-specific culture stratification instead total cultures were used without exclusive disaggregation for confirmed urine specimens.
(iii) No molecular diagnostics with genotypic AMR confirmation e.g., ESBL or carbapenemase gene detection.
(iv) AST testing was not uniformly reported across all drug–pathogen pairs, we focused only on the WHO priority bacterial pathogens.
(v) Several genera data (e.g., *Streptococcus*, *Enterococcus*) lacked precise species identification, limiting all pathogen-specific interpretation.

Despite these limitations, our study remains a critical contribution to global AMR surveillance by providing one of the largest multi-year laboratory-based datasets on uropathogens in Nigeria, offering valuable empirical evidence of resistance gradients and treatment failure threats. The scale, temporal scope, and organism-specific analysis bridge significant knowledge gaps in regional AMR epidemiology, particularly for WHO priority bacterial pathogens like *Klebsiella*, *E. coli*, and *Pseudomonas*. Our findings deliver actionable novel insights into antimicrobial policy revisions, reinforcing the urgency of laboratory strengthening, and create a foundation for more targeted stewardship interventions. This evidence base is pivotal for aligning Nigerian microbiology practices with global AMR containment strategies.

## Conclusions

High AMR across key drug classes for uropathogens like K*lebsiella, Escherichia coli* and *Staphylococcus species* suggests a narrowing window for effective treatment of UTIs in Nigeria. Our data offer a timely, actionable microbiological framework to support diagnostic stewardship, inform empiric treatment updates and guide national AMR containment policies in this region. Translationally, these results emphasize the need for enhanced AMR surveillance, strengthening molecular diagnostics capacity and feedback systems for uropathogens within Nigeria’s microbiology infrastructure. Future perspectives include embedding these new insights into regional and global AMR strategies to strengthen UTIs management and antimicrobial stewardship.

## Data Availability

Data is available as a Supplementary Material

https://aslm.org/wp-content/uploads/2023/07/AMR_REPORT_NIGERIA.pdf?x89467

## Acknowledgments

We would like to thank and acknowledge the Mapping Antimicrobial Resistance and Antimicrobial Use Partnership (MAAP) consortium for enhancing global health research by facilitating the public access of all national AMR surveillance reports, including the Nigerian AMR report which was the source of our data collection. We also appreciate the contributions of Nigeria Federal Ministry of Health, the Fleming Fund Country Grant (Phase 1) team, national AMR coordinators, microbiologists, and data managers who ensured quality-assured reporting across surveillance sites in the report.

## Ethics statement

This study utilized publicly available, de-identified AMR surveillance data originally collected by the Nigerian Ministry of Health under the Fleming Fund Regional Grant (Phase 1, https://aslm.org/wp-content/uploads/2023/07/AMR_REPORT_NIGERIA.pdf?x89467). No individual-level data were accessed. Secondary data use complied with the Declaration of Helsinki and did not require additional ethical clearance.

